# Geographical inequalities in the evolution of the COVID-19 pandemic: An ecological study of inequalities in mortality in the first wave and the effects of the first national lockdown in England

**DOI:** 10.1101/2021.10.23.21265415

**Authors:** Claire E. Welsh, Viviana Albani, Fiona E. Matthews, Clare Bambra

## Abstract

**Objectives:** This is the first study to examine how geographical inequalities in COVID-19 mortality rates evolved in England, and whether the first national lockdown modified them. This analysis provides important lessons to inform public health planning to reduce inequalities in any future pandemics.

**Design:** Longitudinal ecological study

**Setting:** 307 Lower-tier local authorities in England

**Primary outcome measure:** Age-standardised COVID-19 mortality rates by local authority and decile of index of multiple deprivation.

**Results:** Local authorities that started recording COVID-19 deaths earlier tended to be more deprived, and more deprived authorities saw faster increases in their death rates. By 2020-04-06 (week 15, the time the March 23^rd^ lockdown could have begun affecting deaths) the cumulative death rate in local authorities in the two most deprived deciles of IMD was 54% higher than the rate in the two least deprived deciles. By 2020-07-04 (week 27), this gap had narrowed to 29%. Thus, inequalities in mortality rates by decile of deprivation persisted throughout the first wave, but reduced somewhat during the lockdown.

**Conclusions:** This study found significant differences in the dynamics of COVID-19 mortality at the local authority level, resulting in inequalities in cumulative mortality rates during the first wave of the pandemic. The first lockdown in England was fairly strict – and the study found that it particularly benefited those living in the more deprived local authorities. Care should be taken to implement lockdowns early enough, in the right places - and at a sufficiently strict level-to maximally benefit all communities, and reduce inequalities.

**Strengths and limitations of this study:**

- This study interrogates the evolution of inequalities in COVID-19 in the first wave of the pandemic in England and the impact of the national lock down.
- National level official (ONS) data used, covering nearly all local authorities in England and including all deaths that made any mention of COVID-19 on death certificates, requiring sensitive data acquisition.
- Age-standardised deaths rates at lower geographies are not available at the time of writing but could lend extra nuance to these findings.
- Ecological study not using individual level data, so unable to examine the individual level risks for covid-19 mortality.

**Summary Box:** *Section 1: What is already known on this subject:* There are cross-sectional estimates of geographical inequalities in the severity of the COVID-19 pandemic in England in terms of cases, hospitalisations and deaths. But these studies have not examined the evolution of the epidemic nor the impact of the national lockdown on inequalities in COVID-19 related mortality.

*Section 2: What this study adds:* This study provides the first analysis of inequalities in the evolution of the pandemic in different English local authorities and the impact of the first national lock down on them. We estimate geographical inequalities by local authority in the evolution of age-standardised COVID-19 mortality during the first wave of the pandemic in England (January to July 2020) and the impact on these inequalities in the cumulative death rates of the first national lockdown. We found that more deprived local authorities started to record COVID-19 deaths earlier, and that their death rates increased faster. Cumulative COVID-19 mortality inequalities during the first wave of the pandemic in England were moderately reduced by first national lockdown.

## Introduction

Since the early days of the SARS-CoV-2 pandemic in 2020, inequalities in case, hospitalisation and death rates have been noted internationally(1–7). The most deprived populations and areas in the USA, Europe and other high-income countries have suffered up to twice the mortality rates of the least deprived sections of society(2,8,9). In addition, inequalities in disease burden have been noted across levels of income, education, employment, sex, age, and especially between different ethnic groups, where people of Black and minority ethnic backgrounds have suffered many more cases (and deaths) than their white counterparts(10). However, the evolution of geographical inequalities in the pandemic over time - and the impact of national lock downs on them – has not previously been examined. This study addresses this evidence gap by providing the first analysis of inequalities in the evolution of the pandemic in different English local authorities and the impact of the first national lock down on them.

Most countries employed national lockdowns of varying duration and severity to mitigate disease spread, alongside social distancing and hygiene-related advice. The factors used to determine when a lockdown should begin or cease were rarely transparent, but most appeared to reduce infection rates to some degree after a lag phase, and saw a rebound of varying size following their release(11– 13). The first confirmed cases of COVID-19 were recorded in England in York in January 2020 and the first death in England was on March 5^th^. From 2020-04-23 until 2020-07-04, a national lockdown was implemented across England. In keeping with many other European countries, this was characterised by a 12 week ‘stay at home’ order (SI 350) - whereby people could only go outside for certain “very limited purposes” - to buy food, to exercise once a day, for medical reasons or to care for a vulnerable person, or to go to work if they absolutely could not work from home(12). Face-to-face education was suspended and many workplaces closed down - and staff furloughed -particularly in the hospitality, travel and retail sectors. As nationally cases, hospitalisation and death rates started to fall the lockdown was gradually released over a period of several months - culminating in the so-called ‘Super Saturday’ on 2020-07-04 when pubs, restaurants, hairdressers, and cinemas reopened – albeit with strict social distancing rules(13).

It has been noted that when national epidemic dynamics are used to examine population health, they can mask important sub-national variation in disease spread, thus mitigation strategies that rely solely on the national data to inform implementation timings could inadvertently worsen health inequalities across geographical areas(11,13). Previous descriptive studies and reports of inequalities in COVID-19 mortality have only focused on cumulative measures over set timespans, without documenting the disparities in evolution of mortality rates(5,14,15), have been restricted to higher geographies(18), or have not focussed on the effects of lockdowns (7,19). An understanding of how the evolution of the pandemic differed by area and the impact of national mitigation strategies on geographical inequalities in COVID-19 mortality could help inform future policies targeted at minimising viral spread whilst preventing the widening (or even actively decreasing) health inequalities.

This paper uses COVID-19 mortality data from the first wave of the pandemic in England to provide the first interrogation of geographical inequalities in the evolution of the pandemic. It sets out the first analysis of when death rates rose, peaked and fell in local authorities of differing levels of deprivation, and it describes the effects – and the timing of - the first national lockdown on these inequalities.

## Methods

Weekly counts of COVID-19 deaths (based on any mention of Coronavirus on the death certificate) for 312 lower-tier local authorities (excluding county councils) in England were obtained from the Office for National Statistics (ONS) covering the period from 1^st^ January 2020 to 4^th^ July 2020, by date of registration (16). Weekly COVID-19 death counts at the local authority level were not available per age group, thus age-standardised rates were calculated via monthly age-standardised rates. Monthly age-standardised COVID-19 mortality rates per local authority for the period March to July 2020 were similarly obtained from ONS(21). The monthly rate was divided between the constituent weeks based on the share of monthly deaths in each week. Where all age-standardised rates for a local authority were suppressed by ONS due to disclosure controls, the authority was excluded from analyses (n=4). The level of deprivation of each local authority was determined by the rank of average rank of the Index of Multiple Deprivation (IMD), which was converted into deciles (decile 1 contained the most deprived 10% of local authorities) from downloaded data(17). In addition, data from the Isles of Scilly and the City of London were excluded due to well-known mortality data quality issues and low population counts.

A number of metrics were calculated for each local authority; the ‘starting week’ was the first week where 1 or more COVID-19 deaths were registered, the ‘peak’ was the highest weekly age-standardised mortality rate per area using a 3-week rolling mean of weekly death rates, and the ‘total mortality rate’ was the cumulative sum of age-standardised weekly mortality rates over the whole study period. The speed of increase was defined as the change in mortality rate between 25% of peak and the peak rate, divided by the number of weeks between them, and similarly the speed of descent was calculated using the peak rate and subsequent reduction to 50% of peak (25 and 50% selected to include time window when epidemic peaks were visibly most stable). An assumption was made that any change in population incidence of COVID-19 cases may begin to be seen 2 weeks later in mortality data, thus analyses of the effect of lockdown focused on the period before or after week 15 (lockdown was announced in week 13 [March 2020] and ended on ‘Super Saturday’ [July 4^th^, week 27], which is shown in timeline plots). The ‘peak difference’ was the difference in weeks between the peak mortality rate and the week in which lockdown began to take effect (week 15).

Weekly age-standardised mortality rates per IMD decile were not available at the time of writing, thus they were calculated from other existing data, in a similar but distinct method from local authority rates. Firstly, the denominators from local authority-level monthly age standardised mortality rates were calculated using the death counts and rates provided. These ‘modified’ population estimates were summed across local authorities within the same IMD decile, and counts of COVID-19 deaths were similarly summed by decile. Weekly age-standardised rates per 100,000 people were then calculated as the sum of deaths divided by the modified summed population estimate, multiplied by 100,000.

Simple linear models were employed to analyse the associations between visually normally distributed measures such as the total cumulative mortality rate with other metrics and IMD decile. No model selection was employed, covariate inclusion was based on empirical knowledge.

Maps were drawn based on 2020 geographical boundaries from the ONS Open Geography Portal(18).

All analyses were conducted in R statistical software version 3.6.2.

### Patient and Public Involvement

Our public involvement panel inputted into project design and considered the research topic to be of contemporary importance and value. The data used do not require patient permissions for use and are publicly available.

## Results

All 307 lower-tier local authorities in England began registering deaths involving COVID-19 between weeks 11 and 15. The proportion of areas of each IMD decile per ‘starting week’ is shown in Figure 1. From this it can be seen that more deprived areas (most deprived decile = 1) tended to begin recording COVID-19 deaths earlier than less deprived areas (least deprived decile = 10).

**Figure 1.**
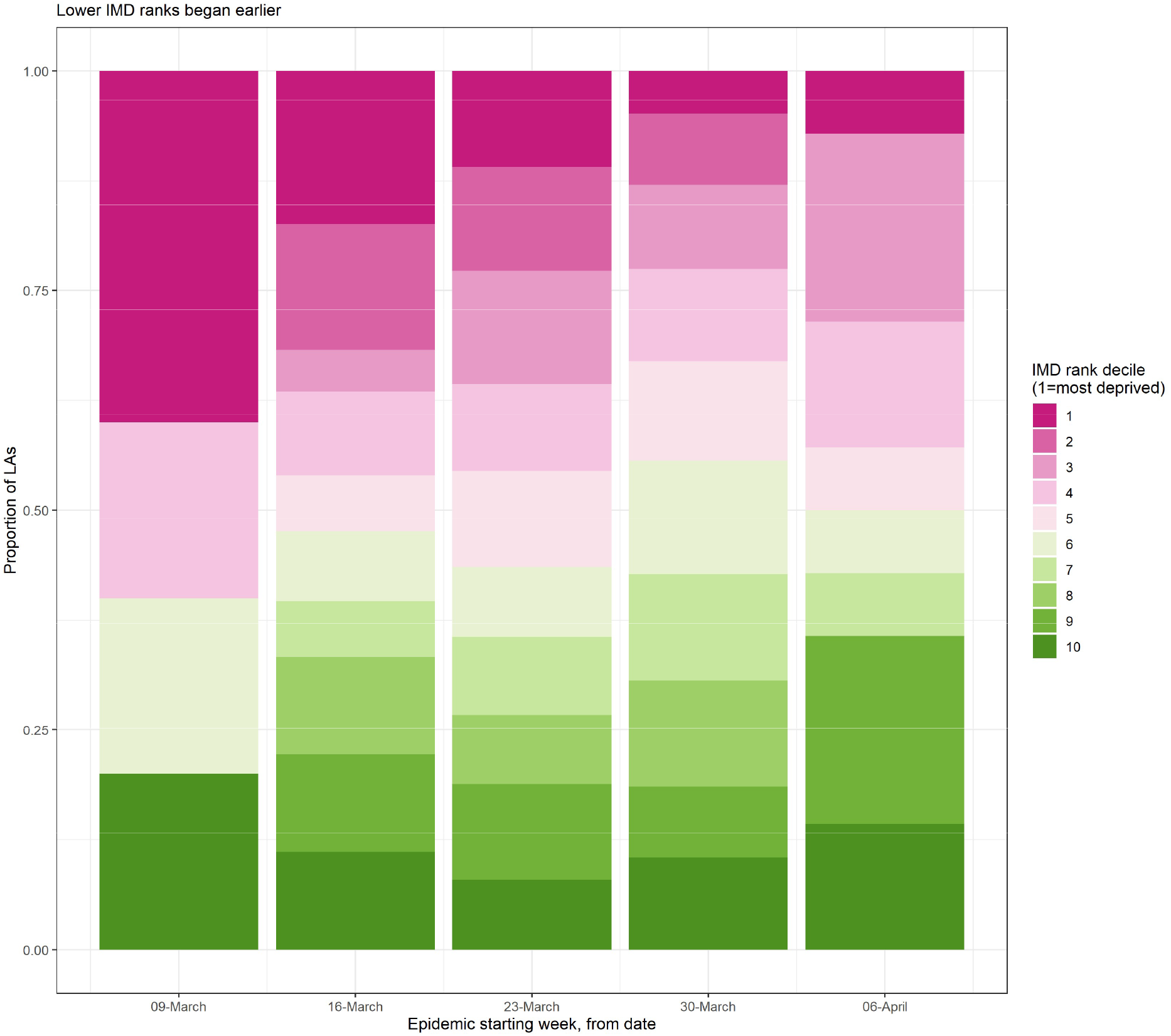
Proportion of 312 English local authorities within each IMD decile that began recording COVID-19 deaths between weeks 11 and 15 of 2020.

Figure 2 depicts the weekly mortality rates per 100,000 people for each IMD decile. After the first two weeks of the epidemic, the two most deprived deciles (20% of local authorities) had the highest speed of increase in age-standardised mortality rates and reached higher peak rates than less deprived areas.

**Figure 2.**
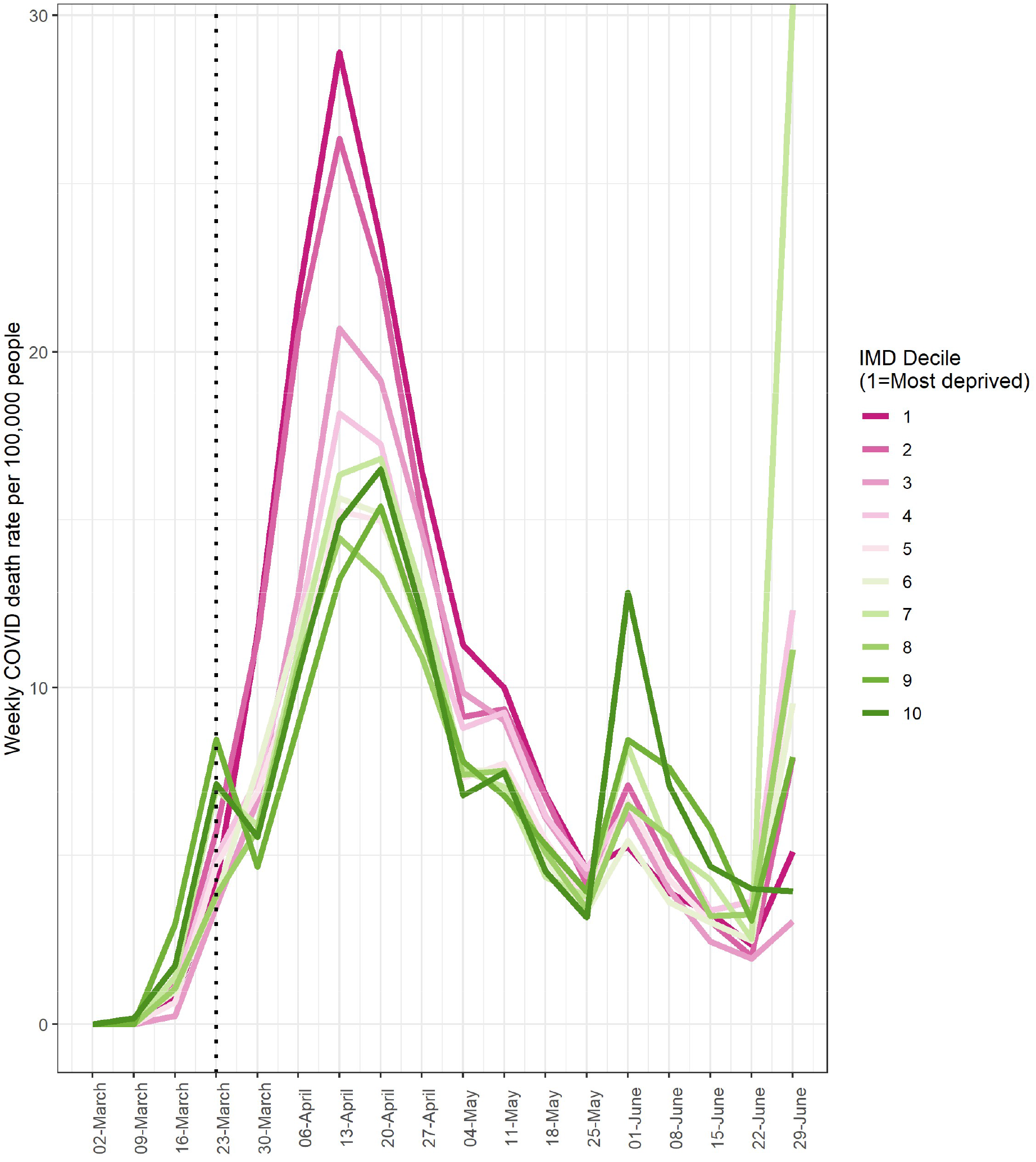
Weekly age-standardised COVID-19 mortality rates per 100,000 in areas of each IMD decile. Dotted line indicates the start of the first national lockdown (26^th^ March).

From the week of their first COVID-19 deaths to week 15 (when lockdown could plausibly have begun affecting death rates), local authorities in the two most deprived deciles had the highest speed of increase in death rate (albeit not statistically significantly different), and the less deprived deciles increased more slowly (Figure 3). The mean speed of increase in two the most deprived local authorities was 4.03 deaths per 100,000 persons per week, and in the two least deprived local authorities was 2.18 deaths per 100,000 persons per week (a difference of 46%).

**Figure 3.**
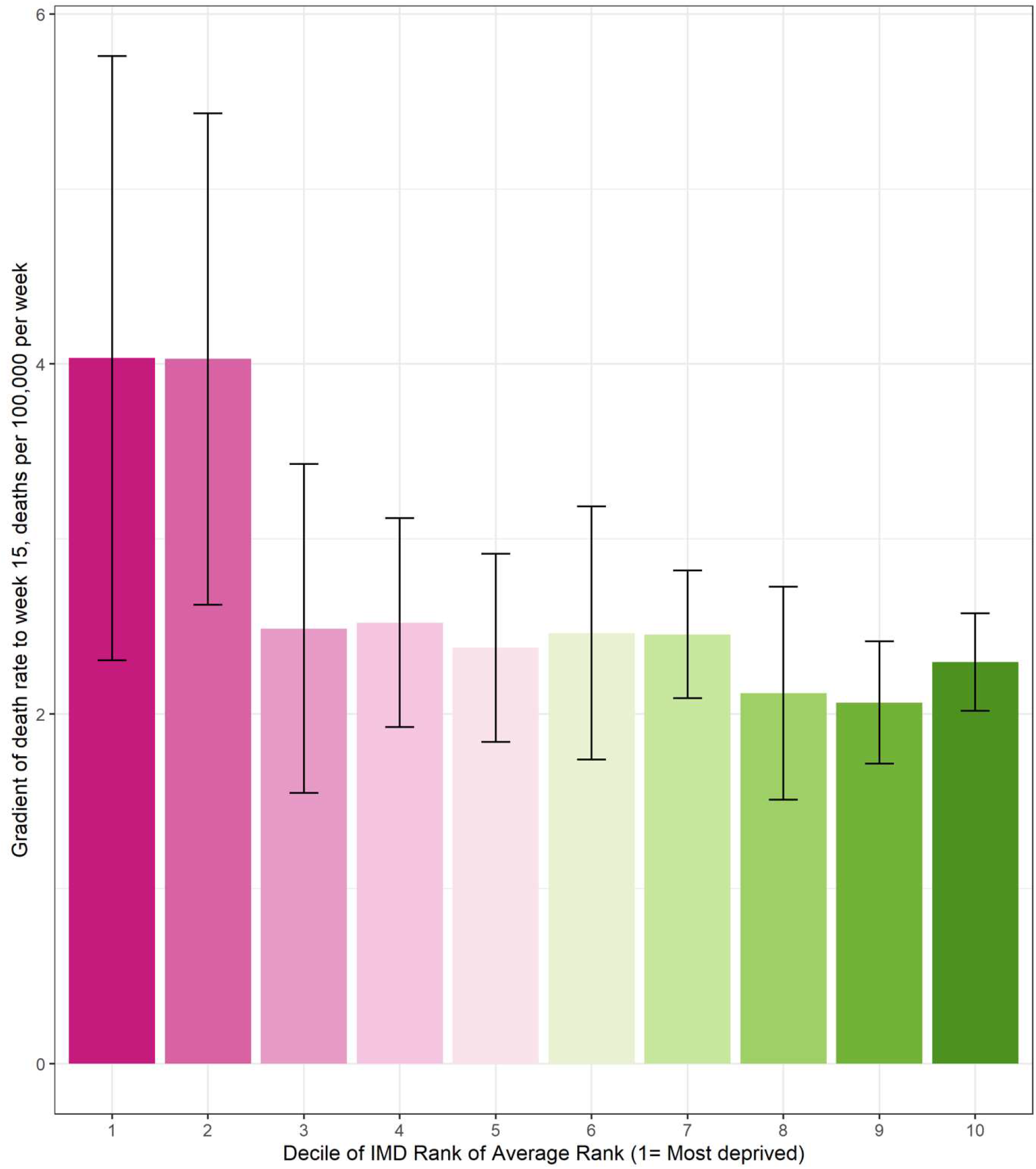
Simple linear gradient of age-standardised COVID-19 death rate per 100,000 people between the first week of recorded COVID-19 deaths and week 15, across rank of average rank of IMD deciles.

All local authorities’ death rate curves peaked and began to decline between 3 and 10 weeks following the start of the first lockdown. Those local authorities whose death rates were increasing faster before lockdown peaked sooner after lockdown commenced compared to slower local authorities.

The total age-standardised cumulative mortality over the first wave (up to week 27, week commencing 2020-06-28) varied from 119 to 2349 deaths per 100,000 persons per local authority. Table 1 describes the multivariable linear model of total cumulative death rates per local authority. It shows that, compared to the most deprived 10% of local authorities, less deprived areas (deciles 3-10) recorded lower cumulative death rates, and that areas with higher speeds of increase - and more weeks of recorded COVID-19 deaths before lockdown (plus those that peaked later) - saw higher total death rates.

**Table 1.**
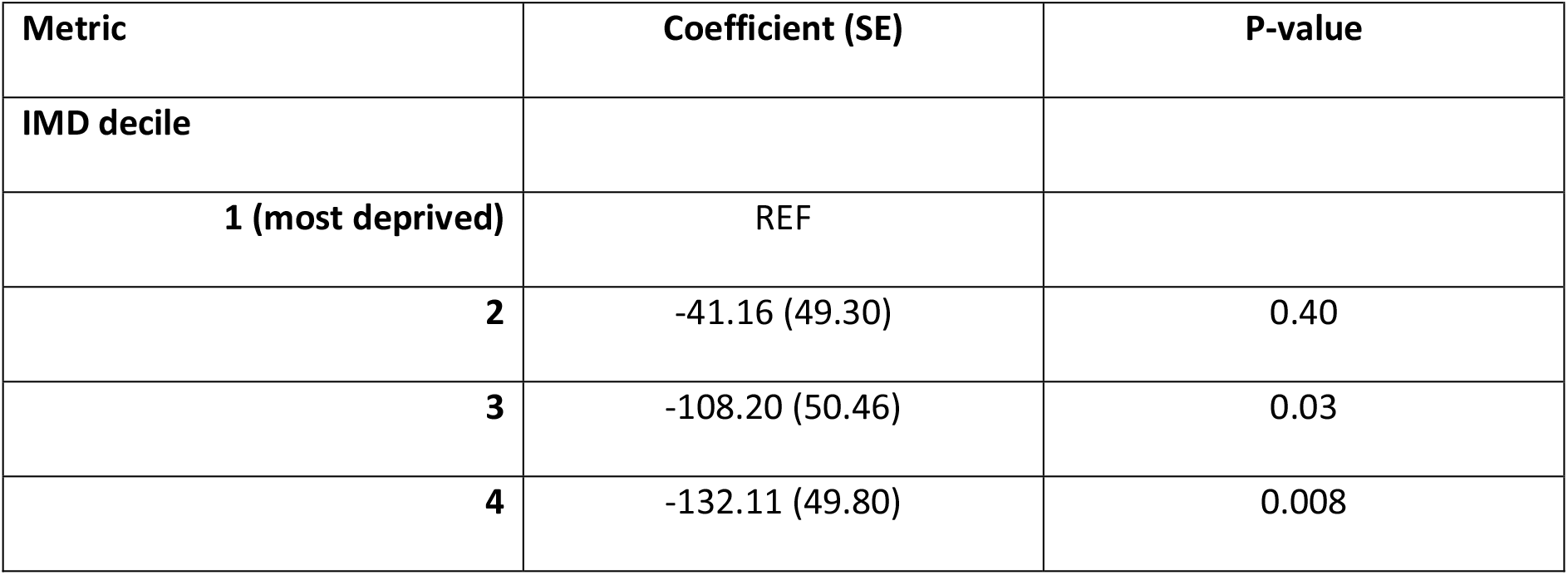
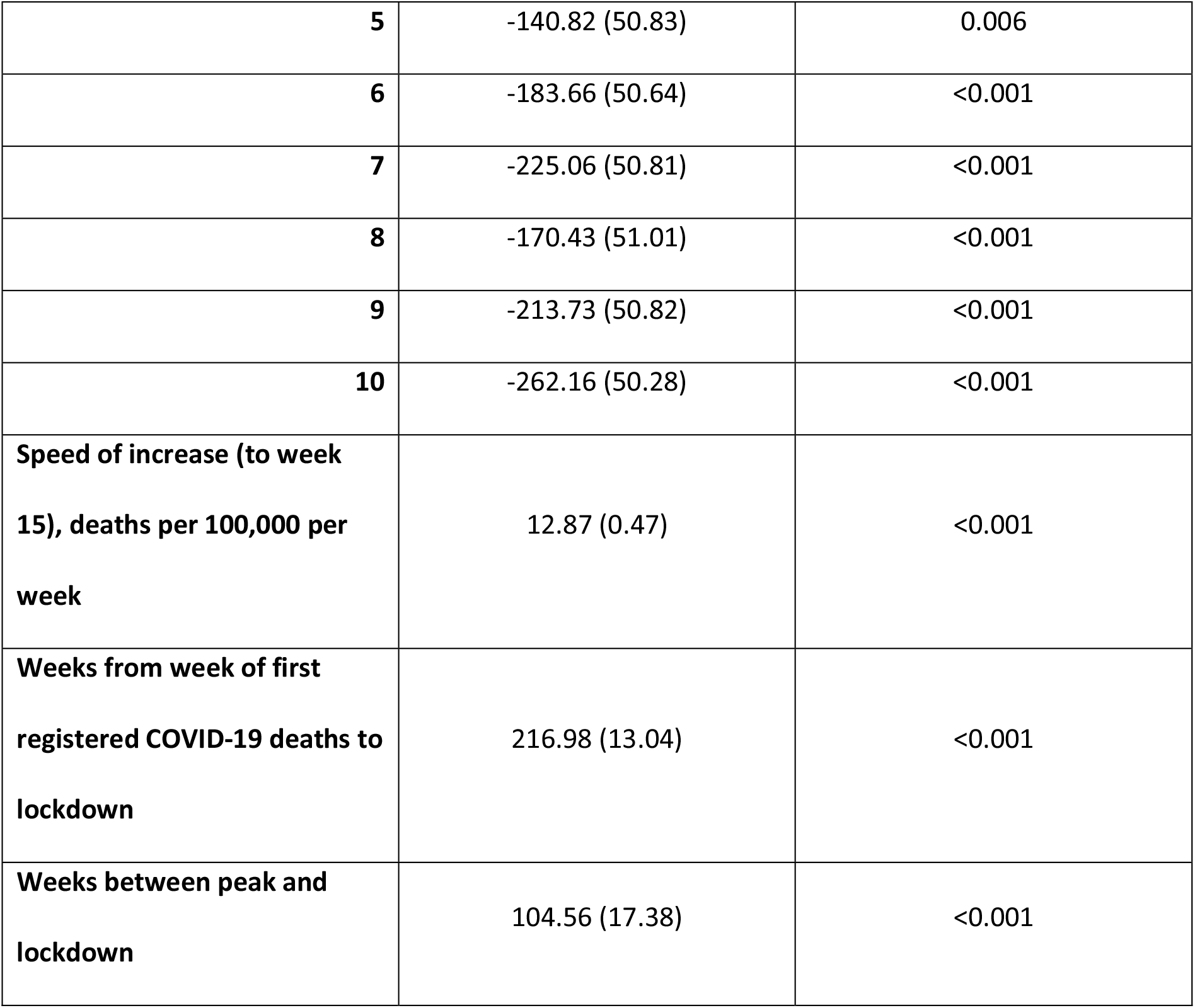
Linear multivariable model of the total cumulative age-standardised COVID-19 death rate per 100,000 persons between weeks 1 and 27 of 2020, among 307 local authorities in England.

As mentioned, all local authorities began recording COVID-19 deaths between weeks 11 and 15, i.e., from 2 weeks before the announcement of the first lockdown, to 2 weeks after. The difference in total cumulative death rates for areas grouped by starting week are as seen in Table 2.

**Table 2.** Mean cumulative COVID-19 death rate per 100,000 persons over the first wave (weeks 1 to 27, 2020) of the pandemic among 307 local authorities in England.

|  |  |  |
| --- | --- | --- |
| **Timing of start week relative to week 13** | **Total cumulative age-standardised COVID-19 death rate per 100,000** | **Number of local authorities** |

| **(when lockdown 1 was announced)** | **persons for whole of wave 1 (weeks 1 to 27, 2020), (SD)** |  |
| --- | --- | --- |
| **2 weeks before** | 465 (451) | 14 |
| **1 week before** | 780 (324) | 124 |
| **Same week** | 984 (407) | 101 |
| **1 week after** | 1188 (505) | 63 |
| **2 weeks after** | 1147 (255) | 5 |

Figure 4 depicts the cumulative COVID-19 death rates of each IMD decile over the whole of the first wave. Mortality rates in more deprived areas (deciles 1 and 2) were rising faster than others at the start of lockdown (vertical dotted line), and the disparity in cumulative mortality grew as the pandemic progressed.

**Figure 4.**
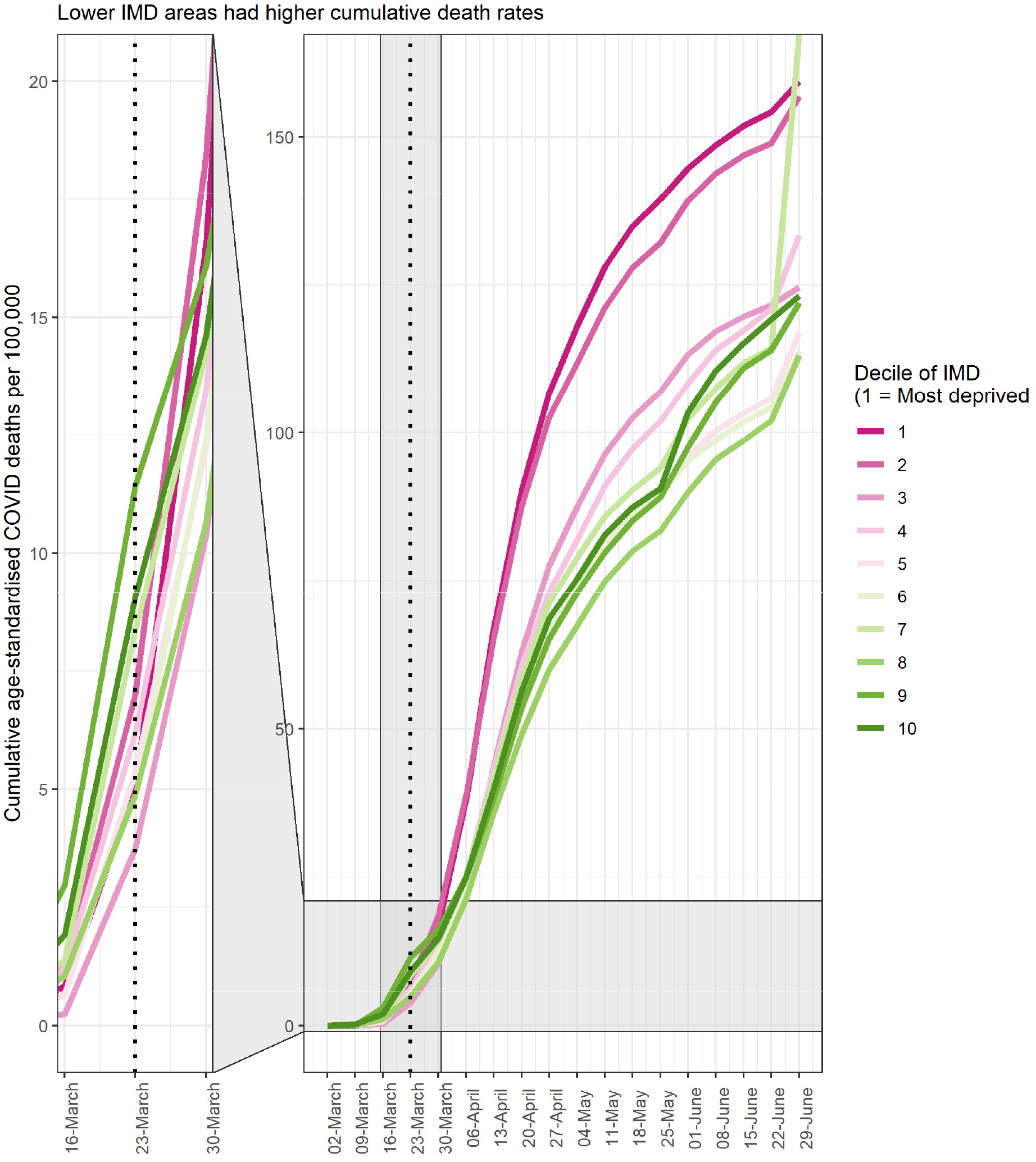
Cumulative COVID-19 death rates per 100,000 for areas of each IMD decile over the first wave of the pandemic in 307 local authorities in England. Dotted line marks timing of the announcement of the first lockdown, zoomed in area between weeks 13 and 14.

Up until week 15 when the effects of lockdown may have started to be seen in mortality data, the cumulative death rate per 100,000 persons already differed by IMD decile. The two most deprived deciles recorded 77.16 deaths per 100,000 persons by this time, whereas the two least deprived deciles recorded only 50.01 deaths per 100,000 persons. This inequality reduced by the time the first wave had passed (by week 27), but did not equalise, with the most deprived two deciles recording 316.14 total deaths per 100,000 persons, and the least deprived recording 245.10 deaths per 100,00 persons. These equate to an excess of 54% before lockdown versus 29% after lockdown.

Figure 5 illustrates the geographical distribution of deprivation based on IMD and the total cumulative age-standardised COVID-19 death rate per 100,000 persons over the first wave of the pandemic. London and the North West featured many of the areas with the highest overall death rates. Although these areas featured many deprived local authorities, the distributions were not identical.

**Figure 5.**
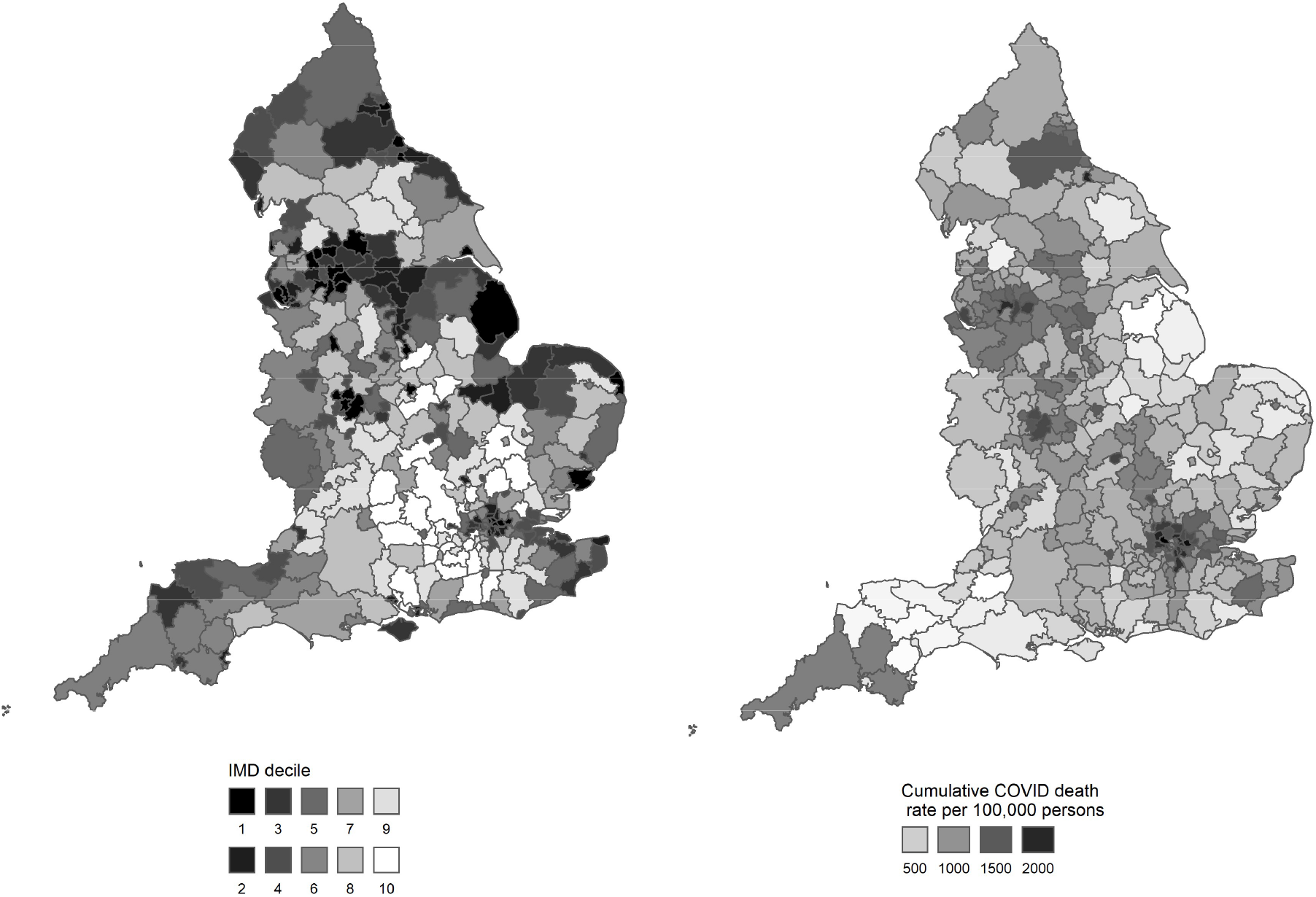
Average rank of the index of multiple deprivation (IMD) and total cumulative COVID-19 death rate per 100,000 persons over the first wave of the pandemic (weeks 1 to 27, 2020) per local authority in England.

## Discussion

This study has provided the first examination of the evolution of inequalities in the COVID-19 pandemic. It has found that inequalities in COVID-19 mortality rates by deprivation in England began to appear early in the first wave. More deprived local authorities generally started recording COVID-19 deaths earlier than less deprived areas, and mortality rates also increased faster in more deprived areas, and rose to higher peak rates. All of the 307 lower-tier local authorities in England began recording COVID-19 deaths as early as 2 weeks before first national lockdown in England was announced, or up to 2 weeks afterwards, with the latter – less deprived - group of local authorities recording fewer cumulative deaths over the whole of the first wave, compared to the former – more deprived – group of local authorities.

The study has also provided the first assessment of the impacts of the first English national lock down on the evolution of the pandemic. It has found that following the implementation of the national lockdown, local authorities where death rates had been rising faster (i.e. more deprived areas), peaked and began to descend earlier than the other – less deprived – local authorities. Cumulative death rates were higher in more deprived areas by the time lockdown began, but the difference narrowed moderately towards the end of the first wave.

England imposed a national lockdown during the first wave of the COVID-19 epidemic in March 2020(19). This measure aimed to drastically reduce instances of interpersonal contact between infected individuals (whether symptomatic or not) and the wider susceptible population. Confining the public to their homes, suspending face-to-face education and restricting travel placed great burdens upon the health and welfare of many individuals and communities, through a number of pathways that are still being elucidated, and which will continue to emerge(20–22). There is no doubt that the economic implications of such lockdowns can be severe, and disruptions to usual health care provision have led to increased mortality from non-COVID causes (23). However, the risks posed to society of not imposing such lockdowns are likely much greater(24). Unchecked viral spread would lead to mass fatalities, increased disability rates especially in the young from the effects of non-fatal infection (so-called ‘Long COVID’(25)), and an increased risk of viral mutation into forms which may pose even greater threat(26). Importantly, the National Health Service (NHS) could potentially be filled beyond capacity with COVID-19 patients, leaving insufficient resources for non-COVID patients of all ages and diagnoses. Economic implications of unchecked viral spread are likely to be considerably worse than those caused by national lockdowns, and could continue for longer due to the likelihood of future outbreaks of mutated viral strains and multiple waves of infection(24). A well-timed national lockdown has the ability to reduce case incidence to low levels at which ‘test, trace and isolate’ programs can efficiently extinguish local outbreaks, and lends time for mass vaccination to offer protection, especially to the most vulnerable. However, a lockdown that is imposed too late, i.e. when disease incidence is already high and rising, needs to be substantially more stringent and protracted to offer the same slowing effect on case numbers and, subsequently, deaths(24).

Previous work has focused on comparing COVID-19 mortality rates between areas of England using set time periods without considering the evolution of the inequalities reported(21), or have identified inequalities in case rates and other metrics(13). Using mortality data removes some of the uncertainty surrounding early case ascertainment, since early in the English epidemic, testing was only being performed in hospitals on symptomatic individuals, and so many infections would not have been recorded.

It has been noted internationally that the seeding of SARS-CoV-2 into a country tends to be via travel by people at the upper end of the socio-economic spectrum, taking international holidays or travelling for business(27,28). Cases then increase within these less deprived populations until social distancing and national lockdowns are advised or mandated. At this point, the disease burden shifts to the more deprived, who are less able to fully adhere to these guidelines due to les ability to work from home, fewer resources, precarious work, higher population densities and other pre-existing factors(27). These two ‘phases’ of pandemic spread likely apply to COVID-19 cases in England, where the index cases were holidaymakers returning from skiing trips to Austria(29,30). Plümper *et al* (2020) reported that in Germany, despite a somewhat reduced likelihood of infection for those in more deprived areas in the first phase of the epidemic, these communities were nevertheless at similar risk of death. This relative risk of mortality increases for more deprived areas once transmission is established in ‘phase 2’ of the pandemic – due to population vulnerabilities including poverty, overcrowding and pre-existing chronic conditions(6). Our analysis of early-stage mortality in England confirmed this structure, in that mortality rates rose first to a small initial ‘peak’ in less deprived areas, before being dominated by more deprived local authorities. The earliest data available to the German study began more than 2 weeks following the implementation of government lockdowns, whereas the analysis we present here predate the UK lockdown by a number of months, and hence capture the very earliest data available on COVID-19 deaths.

We have shown that inequalities in cumulative death rates during the first wave of infection in England existed from the earliest stages of COVID-19 mortality reporting, and were entrenched by differences in the speed of increase, leading to unequal burdens of cumulative mortality at local authority level by the time the first national lockdown was called. These inequalities reduced marginally but were not abolished by the national control measures implemented in the lockdown. The first national lockdown in England was fairly strict (e.g. a ‘stay at home order’) and it was a universal intervention, enforced and applied to the whole population and thereby requiring little by way of individual agency. Previous public health research has shown that such measures are more likely to reduce inequalities in health than those that require individual choice/compliance(31). That the lockdown did not completely eliminate geographical inequalities in COVID-19 mortality may well be as a result of inequalities in (1) *vulnerability* (whereby more deprived areas had a higher burden of clinical risk factors); (2) *susceptibility* (whereby immune response was lower in more deprived populations due to the adverse consequences of long term exposures to harmful living and environmental conditions); (3) *exposure* (inequalities in working conditions notably less ability to work at home in the low income jobs predominating within more deprived local authorities); and (4) *transmission* (higher rates of overcrowding and population density in the community may have impacted on infection spread in more deprived areas)(6).

## Conclusion

This study has found that inequalities in death rates during the first wave of infection in England existed from the earliest stages of the COVID-19 pandemic, and were entrenched by differences in the speed of increase. This led to a significant unequal burden in cumulative mortality between the most and least deprived local authorities by the time the first national lockdown was implemented. These inequalities reduced marginally - but were not abolished - during the national lockdown. It is impossible to say with certainty whether an earlier – or longer - national lockdown could have further reduced these inequalities, but it should be noted that, although the lockdown did reverse the trend in mortality rates across the country, it had to do so at more advanced stages of the epidemic in more deprived areas, compounding the unequal disease burden upon these communities and local health care systems. Susceptibility to infection and fatality from COVID-19 is undoubtedly closely associated with deprivation, but other factors also play an important part, as well as the stochasticity implicit in viral spread. Nevertheless, our understanding of how deprivation associates with mortality from a novel infectious disease within a virgin population it can help to focus future public health attention on those communities most in need and at risk.

## Limitations

Weekly age-standardised mortality rates were not available at local authority level at the time of writing. However, we were able to pro rata monthly age-standardised rates to weekly ones using weekly death counts. Age-standardised weekly rates are unlikely to become available at lower geography levels due to disclosure risks. Death counts did not include deaths of non-residents of England, nor where place of residence was unknown, and was based on date of registration rather than date of death.

Deprivation is undoubtedly linked to COVID-19 mortality, it cannot explain all of the variation in area-level mortality rates, hence COVID-19 mortality and IMD are not perfectly correlated. Many other factors including comorbidity, healthcare provision, employment types and variation in transport links all likely play a part in the causal web linking lockdowns to mortality inequalities. A deeper analysis of these underlying associations was beyond the scope of the current paper, but warrants further scrutiny.

## Data Availability

All data produced in the present study are available upon reasonable request to the authors

## Authorship

FM, CB and CW designed the study. CW completed all analyses with input from FM and CB. CW, VA, FM and CB all contributed to drafting the manuscript. The corresponding author attests that all listed authors meet authorship criteria and that no others meeting the criteria have been omitted. CW is guarantor of the analysis.

The Corresponding Author has the right to grant on behalf of all authors and does grant on behalf of all authors, a worldwide licence to the Publishers and its licensees in perpetuity, in all forms, formats and media (whether known now or created in the future), to i) publish, reproduce, distribute, display and store the Contribution, ii) translate the Contribution into other languages, create adaptations, reprints, include within collections and create summaries, extracts and/or, abstracts of the Contribution, iii) create any other derivative work(s) based on the Contribution, iv) to exploit all subsidiary rights in the Contribution, v) the inclusion of electronic links from the Contribution to third party material where-ever it may be located; and, vi) licence any third party to do any or all of the above.

## Data Sharing Statement

All data used are publicly freely available through the ONS. Code used in the analyses is available upon request.

## Ethical Approval

This study was approved by the Newcastle University Ethics Committee (Ref: 7543/2020).

## Transparency declaration

The lead author^*^ affirms that this manuscript is an honest, accurate, and transparent account of the study being reported; that no important aspects of the study have been omitted; and that any discrepancies from the study as planned (and, if relevant, registered) have been explained.

*The manuscript’s guarantor.

